# High Seroprevalence to *Aedes*-borne arboviruses in Ethiopia: a Cross-sectional Survey in 2024

**DOI:** 10.1101/2024.09.19.24313917

**Authors:** Daniel M. Parker, Werissaw Haileselassie, Temesgen Sisay Hailemariam, Arsema Workenh, Salle Workineh, Xiaoming Wang, Ming-Chieh Lee, Guiyun Yan

**Affiliations:** Department of Population Health and Disease Prevention; University of California, Irvine; U.S.A; Department of Epidemiology & Biostatistics; University of California, Irvine; U.S.A; School of Public Health, College of Health Sciences; Addis Ababa University; Addis Ababa, Ethiopia; Medical Diagnostic Laboratory, Tikur Anbessa Specialized Hospital; College of Health Sciences, Addis Ababa University; Addis Ababa Ethiopia; Dil chora General Hospital, Dire Dawa Health Bureau, Dire Dawa City, Ethiopia

**Keywords:** dengue, Zika, chikungunya, *Aedes aegypti*, Ethiopia, arbovirus, seroprevalence

## Abstract

*Aedes*-borne diseases infect millions of people each year. In the last decade several arbovirus outbreaks have been reported in Ethiopia. Arbovirus diagnosis and surveillance is lacking and the true burden is unknown. In this study we conducted a seroprevalence survey using a commercially available test kit that that tests for immunological responses to IgM and IgG for dengue (DENV), Zika (ZIKV), and chikungunya (CHIKV) viruses in Dire Dawa city, eastern Ethiopia. We found a high IgG seroprevalence for DENV (76%), CHIKV (44%), and ZIKV (38%), and <20% IgM seropositivity across all viruses. As a comparison, we conducted serosurveillance in Addis Ababa, the national capital with no reported history of arbovirus outbreaks. The highest seropositivity we found was to IgM for DENGV at approximately 3%. Our results suggest both past and recent widespread exposure to these arboviruses, underscoring the need for improved surveillance and public health interventions in Ethiopia.

## BACKGROUND

*Aedes* borne diseases contribute significantly to the global burden of disease, and are especially prevalent in tropical and subtropical settings. *Ae. aegypti*, and to a lesser extent *Ae. albopictus*, are the major vectors of disease and three viruses of primary concern are are dengue virus (DENV, from genus *Orthoflavivirus*), Zika (ZIKV, from genus *Orthoflavivirus*), chikungunya (CHIKV, from genus *Alphavirus*).

DENV, CHIKV, and ZIKV viruses each represent significant public health challenges with distinct impacts and distributions. DENV, endemic in tropical and subtropical regions, is responsible for an estimated 400 million infections annually, with severe cases leading to dengue hemorrhagic fever and dengue shock syndrome^1^. CHIKV was first detected in Tanzania in 1952 and in recent years has caused epidemics in Africa, Asia, Europe, and the Americas^2,3^. The disease is characterized by debilitating joint pain and arthritis that can last weeks to months, with significant economic implications on top of health impacts^4^. ZIKV was first detected in Uganda in 1947 and in the last decades there have been epidemics in Africa, Asia, the Americas, and the Pacific^5^. While most ZIKV infections are mild, the disease has been associated with congenital abnormalities (e.g. microcephaly) when transmitted from mother to fetus^6–8^. Collectively, these mosquito-borne viruses cause substantial morbidity, impact quality of life, and strain healthcare systems globally.

While these viruses cause considerable disease globally, infections are presumptively diagnosed in much of the world, based on symptoms and without laboratory confirmation of the cause of disease. Dengue and chikungunya infections result in similar symptoms and may be confused in some settings in the absence of laboratory confirmation. Zika viruses are often asymptomatic. All three may be drastically under-diagnosed in some settings and surveillance is lacking in much of the African continent so that the true burden of these diseases is unknown.

Seroprevalence surveys offer the possibility to evaluate both the long- and short-term epidemiology of these diseases. Antibodies to IgM typically emerge shortly after infection (normally within days for each virus) and can persist for months. Antibodies to IgG typically emerge weeks after infection and can persist for years (in the case of dengue, may be lifelong). Seroprevalence estimates from IgM are therefore normally indicative of recent epidemiology of these diseases whereas seroprevalence estimates of IgG are indicative of long-term dynamics. In some high transmission settings, most of the population will be seropositive before reaching adulthood^9,10^. Cross-reactivity can complicate seroprevalence surveys that attempt to distinguish exposure to these three viruses^11^.

Several dengue fever outbreaks have been documented in Ethiopia since 2013, with recurrent outbreaks occurring in the Eastern part of the nation^12–15^. Chikungunya outbreaks have also been reported more recently in the Eastern part of the nation^16,17^, and seropositivity among symptomatic individuals has been reported from the Northwest^18^ and more recently in the Northeast part of the nation in patients with suspected malaria infections^19^. Dire Dawa, a major city in the Eastern part of Ethiopia has been repeatedly impacted by dengue infections and more recently by a major chikungunya outbreak that infected over 40,000 individuals^16^. Entomological surveys have confirmed the presence of *Ae. aegypti* in this city as well^20^. Research on Zika infections in this setting is scarce^21^. Surveillance is lacking for all three viruses.

The primary objective of this research were to assess the seropositivity to DENGV, CHIKV, and ZIKV in Dire Dawa, Ethiopia. Since age patterns in seropositivity can reflect both recent and historical transmission intensity, our goal was also to examine potential associations between age and seropositivity. We conducted secondary analyses of cross-reactivity.

## DATA AND METHODS

### Study location

Dire Dawa is a major trade city in Eastern Ethiopia. The city has a hot, arid climate with a hot season from February through June and a two rainy seasons: a major one from July through September and a minor rainy season In March – May. Temperatures normally range from 58F to 91F (14.4C to 32.7C). The wettest month is normally August, with a typical month experiencing a 5 inches (12.7cm) of cumulative precipitation. The estimated population of the city in 2024 is 485,558 and the city is at approximately 1,276 meters elevation.

We conducted a seroprevalence survey in Dire Dawa in March 2024. A total of 340 individuals were recruited for the survey in Dire Dawa. Individuals were selected using a household registry and a table of random numbers, with households being the unit of sampling. In a selected household, all individuals were invited to participate in the survey regardless of age. We specifically targeted kebeles (administrative units) with known dengue outbreaks in previous years for this survey.

For comparison, we conducted a secondary seroprevalence survey on the outskirts of Addis Ababa using the same approach (selecting individuals by household). Addis Ababa is the capital and largest city in Ethiopia, with an estimated population in 2024 of 5,703,628. In contrast to Dire Dawa, Addis Ababa has a mild and temperate climate, rarely experiencing temperatures above 80 F (26.7C) and typically ranging from 48F to 75F (8.9C to 23.9C). The rainy season follows the same general pattern as Dire Dawa, but with August typically experiencing 8 inches (20.3cm) of cumulative precipitation. Addis Ababa is at an elevation of approximately 2,355 meters.

For this secondary survey we recruited a total of 180 individuals to the seroprevalence survey. We hypothesized that exposure to *Ae. aegypti* and associated viruses would be lower in Addis Ababa than in Dire Dawa and wanted to conduct this secondary survey to compare overall seroprevalence results to Dire Dawa, in part because we lacked a diagnostic control. We targeted 2 kebeles in Addis Ababa for this secondary survey.

### Seropositivity test

We used the Chembio DPP^®^ ZCD IgM/IgG system for the seroprevalence survey^22^. This system is an immunochromatographic test, meant for detection and differentiation of IgG and IgM antibodies to DENV, CHIKV, and ZIKV (i.e. each individual potentially testing positive for 6 different outcomes). The test kit comes with a reader, which is used 10 to 15 minutes after adding buffer to the test device.

The reader for the rapid diagnostic test also outputs a numeric value of the intensity of the immunological response. The numeric value corresponds to the intensity of the colored lines on the test device, which are proportional to the presence of antibodies. The reader uses photometry to measure light reflectance from the lines on the test strip, with the intensity of reflected light being inversely proportional to the relevant antibody responses. This quantitative measure provides an opportunity to look at patterns of cross-reactivity across viruses and immunological measures.

### Questionnaire

The survey team visited each household with a tablet that was loaded with basic demographic questions (age and gender) and test responses, and test kits.

### Generalized Estimating Equations

We used a generalized estimating equations (GEE) model to estimate the overall seroprevalence to IgG and IgM for all three viruses, accounting for the household clustering design of the study by specifying an exchangeable correlation structure. We used bootstrapping to obtain robust confidence intervals around the seroprevalence estimates. We then used another GEE model to estimate age-specific seropositivity to IgG and IgM to each of the three viruses (DENGV, CHIKV, and ZIKV). To model the relationship between age and seropositivity, we incorporated natural spline functions, allowing us to capture potential non-linear patterns in seropositivity across age. The model estimated the probability of being seropositive for each virus as a function of age, adjusted for household clustering. We then used these model estimates to generate predicted probabilities of seropositivity across age groups. Confidence intervals for these predictions were calculated by deriving the standard errors of the linear predictor, which were then transformed to the probability scale using the logistic function. These predicted probabilities, along with their 95% confidence intervals, were plotted to visualize how IgG and IgM seropositivity changes with age for each virus, while accounting for clustering at the household level.

### Assessment of cross-reactivity

We used linear mixed models, with the quantitative immunological measures for each of the viruses and for both IgG and IgM as outcome variables, and with the other IgG or IgM measures for each virus as predictors, while accounting for household clustering using a random intercept for household. For models that used IgM measures as the outcome we also included IgG measures as covariates because we hypothesized that previous exposure to other viruses could potential impact immunological reactions to a more recent infection.

### Assessment of household clustering

The use of linear mixed models in our assessment of cross-reactivity also allowed us to investigate household clustering in immunological responses. To quantify household clustering we used the intraclass correlation coefficient (ICC). The ICC quantifies the proportion of variance attributable to between-group differences (in this case, households) compared to within-group differences (individuals). The mixed models were fitted with random intercepts for households, allowing us to estimate the variance in responses across households. By comparing the random intercept variance to the total variance (random intercept variance plus residual variance), we calculated the ICC for each model. This provided insight into how much of the variation in IgG and IgM responses could be attributed to clustering within households versus individual-level variability.

### Ethics review

Ethics approval was obtained from the Institutional Review Board of the College of Health Sciences, Addis Ababa University. Verbal and written informed consent were required from individuals before participating in the study. Assent/consent for participation for those under 18 years old was granted by their parent or legal guardian.

## RESULTS

### Overall seroprevalence

A total of 340 individuals were recruited for the seroprevalence survey in Dire Dawa. Seropositivity to dengue virus IgG was 76% (95% Cis: 0.71 – 0.80). Chikungunya and Zika IgG seropositivity were 44% (0.38 – 0.50) and 38% (0.31 – 0.44), respectively. Seropositivity to IgM antibodies, typically indicating recent infections, was much lower (**Table 1**). Dengue and chikungunya IgM was approximately the same (0.17 (0.13 – 0.21) versus 0.18 (0.13 – 0.22), respectively). Zika IgM was much lower, at approximately 0.03 (0.02 – 0.05).

**Table 1:**
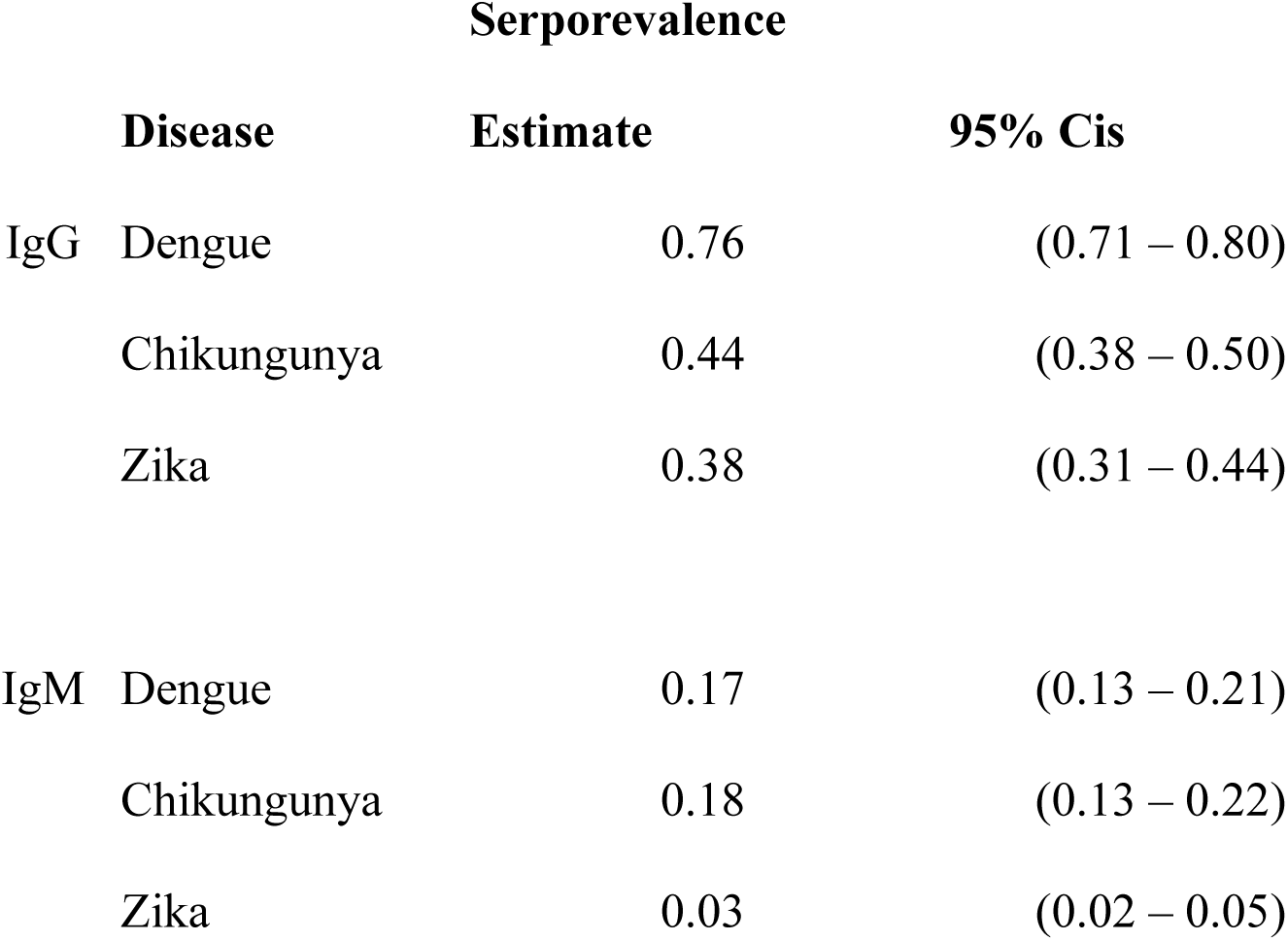
Seropositivity estimates and confidence intervals, from GEE models using an exchangeable correlation structure to account for potential clustering in households.

In comparison, few individuals from the secondary seroprevalence survey in Addis Ababa tested positive (not enough for accurate seroprevalence estimates). A total of 180 individuals were recruited in this survey and out of them 5 tested positive for DENV IgG, 2 for ZIKV IgG, and 1 for CHIKV IgG. Six tested positive for DENV IgM, 1 tested positive for ZIKV IgM, and 1 tested positive for CHIKV IgM. Again, each of the ZIKV IgG positives were also DENV IgG positive. A total of 11 individuals were found seropositive (combining IgG and IgM, and for all three viruses), these individuals ranged in age from 25 to 36 (mean = 30.2; median = 28), and each reported travel to other regions in Ethiopia (five specifically mentioned travel to Dire Dawa).

### Age patterns in seropositivity

Age patterns in predicted probability of seropositivity (**Figure 1**) largely followed the age patterns of the age-specific seropositivity calculations (**Supplemental Figure 1**). By age 10, 52% of all sampled individuals were seropositive to dengue IgG. By age 40, approximately 86% of all sampled individuals were seropositive to dengue IgG. By age 30, over half (51%) of all individuals were seropositive to chikungunya IgG. Finally, by age 20 over 37% of all individuals were seropositive to Zika IgG. The potential cross-over between IgM for dengue and chikungunya was likewise evident in these estimates, though the confidence bands around the estimates were sufficiently large to warrant caution in interpreting this result.

**Figure 1:**
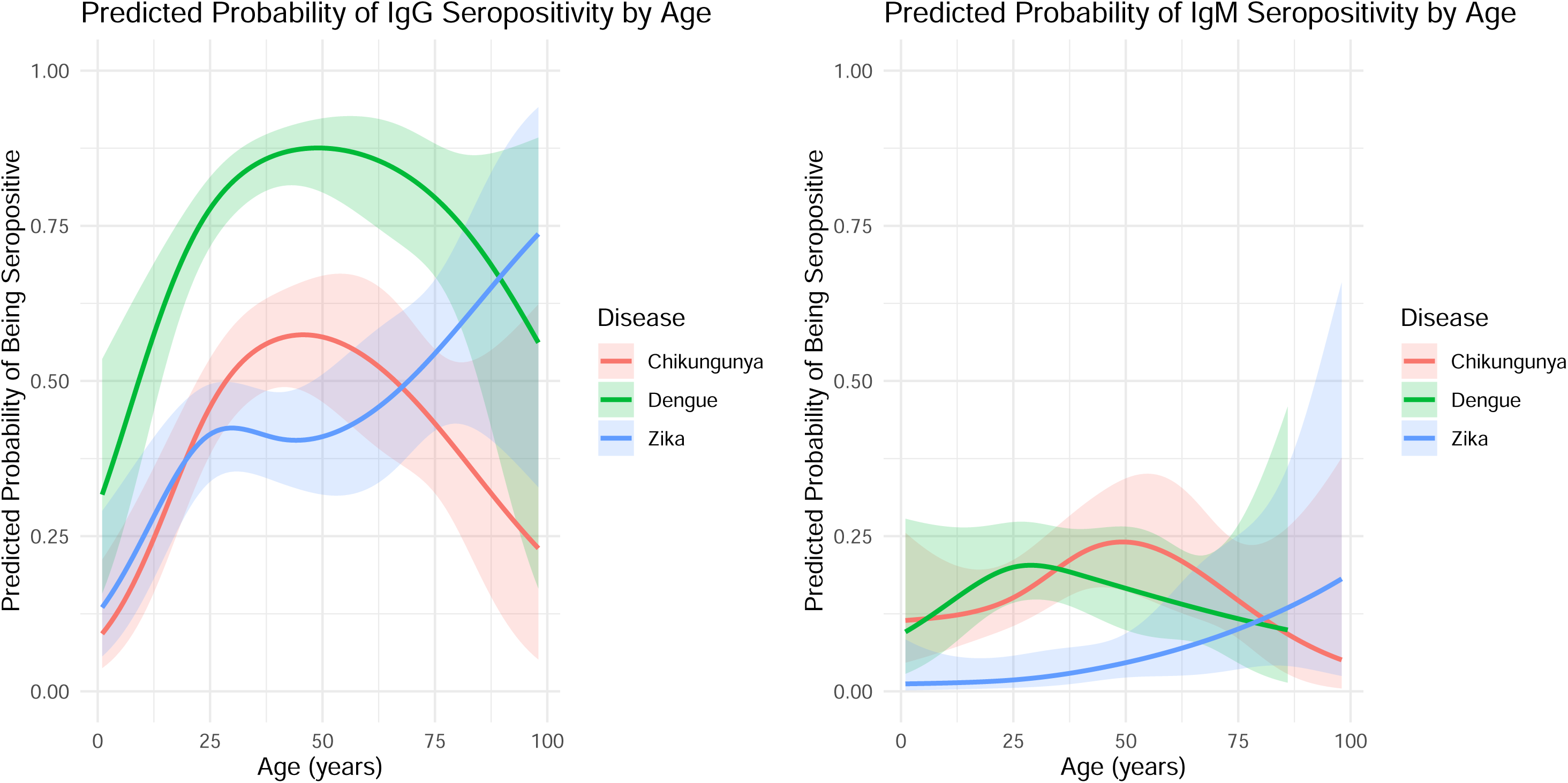
Age specific probability of IgG and IgM seropositivity from the GEE models

### Potential for cross reactivity

All Zika IgG positive individuals were also dengue IgG positive, suggesting a potential for cross-reactivity. Indeed, both are flaviviruses (i.e. genus *Orthoflavivirus*), and so cross-reactivity would not be surprising. 253 of the tested individuals were dengue IgG seropositive and 126 were Zika IgG positive.

An analysis of the quantified immunological response to IgG antibodies for Zika and dengue showed that individuals who were categorized as Zika IgG negative but dengue IgG positive had very low Zika IgG responses. Those who were both Zika IgG and dengue IgG positive had much higher responses than those who were dengue IgG positive but not Zika IgG positive (**Supplemental Figure 2**).

Individuals who were dengue IgG positive and Zika IgG positive had significantly higher quantitative dengue IgG values when compared to those who were dengue IgG reactive but Zika IgG non-reactive (**Supplemental Table 2C**). In general, cross reactivity was high with regard to quantitative dengue IgG outcomes for those who have had a previous Zika exposure. The inverse (cross reactivity where dengue virus exposure increases Zika IgG) appears minimal in comparison. We therefore hypothesize that the Zika IgG positives are not solely the results of cross-reactivity with dengue virus but rather indication of true previous infections with Zika virus. Furthermore, not all Zika IgM positives were also dengue IgM positive.

We generated a Spearman’s correlation matrix from all possible immunological outcomes (**Figure 2**). The strongest associations were between Zika IgG and dengue IgG, chikungunya IgG and chikungunya IgM, followed by Zika IgG and dengue IgM, and dengue IgG and IgM.

**Figure 2:**
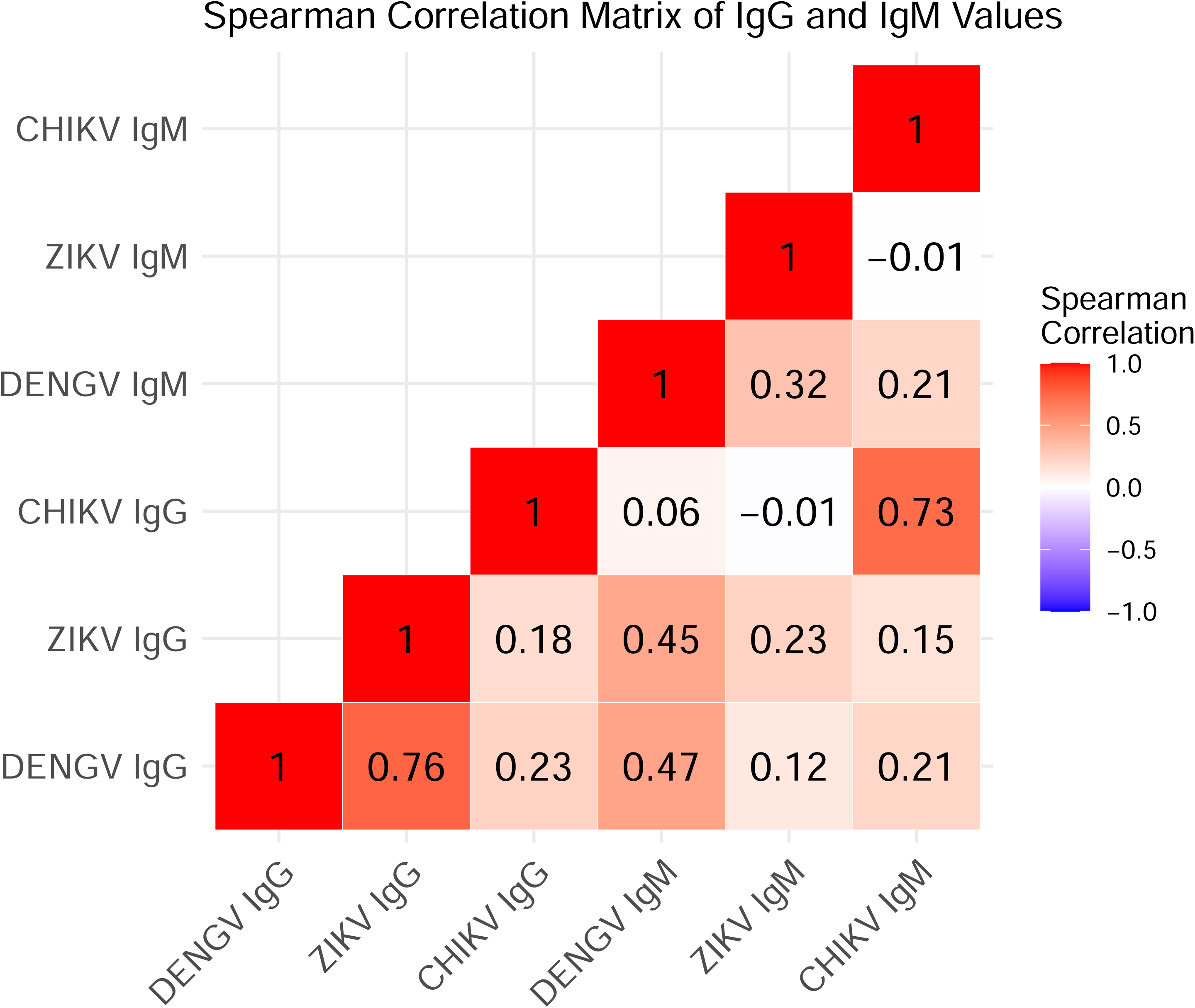
Spearman correlation matrix for correlations between IgG and IgM for each arbovirus

Our linear mixed models analyses (**Supplemental Tables 1 and 2**) showed that Zika IgG was a strong predictor of dengue IgG values (**Supplemental Table 1**). Chikungunya IgG was also associated but the association was smaller than with Zika IgG. Dengue IgG was a strong predictor of Zika IgG. Chikungunya IgG was negatively associated with Zika IgG. Finally, Zika IgG was positively associated with chikungunya IgG values while dengue IgG was negatively associated with chikungunya IgG.

Zika IgM was positively associated with dengue IgM, as was chikungunya IgM (**Supplemental Table 2**). Likewise, dengue and chikungunya IgM values were positively associated with Zika IgM. Finally, dengue and Zika IgM were both positively associated with chikungunya IgM. IgG values were statistically significant predictors of each IgM outcome, but their effect sizes were small.

### Household clustering

The ICC values revealed varying degrees of household clustering for the different viruses and immune responses. For DENGV, we observed moderate clustering in IgG responses (ICC = 10.3%) and a low degree of clustering for IgM responses (ICC = 1.8%). CHIKV showed a similar pattern, with clustering in IgG (ICC = 7.1%) and a stronger clustering effect in IgM responses (ICC = 14.0%). Zika, on the other hand, displayed no household-level clustering for either IgG or IgM responses (ICC = 0 for both). These results suggest that household membership affected the variability in DENGV and CHIKV responses to a moderate extent, while it had no impact on ZIKV responses.

## DISCUSSION

Using a cross-sectional seroprevalence survey from Dire Dawa, Ethiopia we found and here report a high seroprevalence to three arboviruses of concern: DENV, CHIKV, and ZIKV. These results existed for both IgG and IgM suggesting both a history of widespread exposure to these viruses, and recent exposure. The age patterns for DENV and CHIKV IgG seroprevalence in particular suggest that most of the population in this setting has been exposed to these viruses by age 30 (over 50% seroprevalence to CHIKV IgG and over 80% to DENGV IgG). We also found evidence of household clustering with regard to DENGV and CHIKV exposure, but not ZIKV exposure.

While approximately 25% of all children ages 0 – 4 were seropositive to DENGV IgG, none were seropositive to CHIKV IgG. In 2019 there was a major CHIKV outbreak in this setting, with over 40,000 individuals diagnosed with the disease^16^. It is therefore possible that much of the seroprevalence to CHIKV IgG that we found was the result of this previous epidemic, or to the 2019 epidemic and others. We did, however, find individuals in most ages (but not the youngest age group) who were seropositive to CHIKV IgM, suggesting that there has potentially been some recent transmission of this disease or a related alphavirus, among individuals from the study location. IgM to Chikungunya has been shown to persist in some individuals over long periods of time (28 months post infection^23^) but we believe this is unlikely to explain the IgM seropositivity to CHIKV from our survey (now 5 years after the major 2019 epidemic).

While dengue and chikungunya epidemics have previously been reported in Ethiopia, data on exposure to Zika is scarce. One survey from Gambella (in far West Ethiopia) estimated 2.9, 15.6, and 27.3% seroprevalence for Yellow Fever, chikungunya, and Zika virus^24^. Another report of Zika seropositivity in Ethiopia^21^. Zika virus disease has been reported in traveler to Europe coming from Kenya^25^. Zika virus was originally discovered on the African continent, but surveillance remains low^5^.

One limitation of our work is the use of a single screening kit. It might have been preferable to have another test (e.g. ELISA) for comparison on at least a subsample. A few other studies have used the same test kit in research settings. One study suggested general low sensitivity of this test in comparison to ELISA. However, specificity was high (sensitivity/specificity: Dengue 43.92/100%; Zika 25.86/98.81%; CHIKV 37.78/99.35%). If the same pattern holds true in this setting it would indicate that the estimated seroprevalence is an underestimate^26^. Another study focusing on IgM alone found sensitivity/specificity of the ZCD test to Zika IgM was 79.0/97.1%; dengue was 90.0/89.2%; and for chikungunya it was 90.6/97.2%^27^. We take the results of these previous studies to suggest that for ZIKV and CHIKV in particular, our results may be underestimates.

Cross reactivity may have influenced some of our results. DENGV and ZIKV are both flaviviruses, and cross-reactivity between the two is well-known but not completely understood^11,28,29^. If other alphaviruses are circulating in this setting, they could potentially lead to false-positive CHIKV seropositivity. Cross-reactivity can have clinical implications, as is already well-known within DENGV infections across serotypes and has been hypothesized between DENGV and ZIKV infections. While DENGV infections are known to provide long-term immunity, the immunity is sero-type specific and infection by another serotype can lead to more severe disease outcomes (including death) through antibody dependent enhancement (ADE)^30–32^. ADE may also occur across flaviviruses – with the potential for previous ZIKV infection leading to severe dengue disease even in the primary infection by DENGV^33^. Current dengue vaccines are only provided to populations that already have previous exposure to DENGV (i.e. the vaccines should only be provided to seropositive individuals) as vaccinating individuals who are seronegative can increase their risk of severe disease when they are infected by DENGV^34^.

Cross-reactivity is further complicated by the fact that these three viruses are all vectored by the same mosquito (e.g. *Ae. aegypti*), meaning that individuals who have been infected with dengue in this setting are likely to also have been at risk of chikungunya (which has been confirmed in this setting) and Zika virus infection, if the respective viruses are present in, or introduced to, this location. That is, individuals in this setting who acquire dengue infections are likely to also be at risk of CHIKV or ZIKV infections, if those viruses are present, given the shared mosquito vector that is present in this setting.

This seroprevalence survey was meant to look for age-specific seroprevalence in kebeles in Dire Dawa that are known to have previously had dengue outbreaks. While we used a clustered random sampling approach (randomly selecting households within the kebeles), our results may not be representative of the entire city of Dire Dawa. It would be valuable to conduct further surveys that would be representative of the city, as well as in other parts of Ethiopia.

In summary, we found evidence of widespread exposure to DENGV, CHIKV, and ZIKV in Dire Dawa, Ethiopia. This aligns with reported outbreaks of dengue fever in this setting, and of the massive chikungunya fever outbreak in 2019. We also found evidence of exposure to ZIKV, which has rarely been reported from this nation. Not only did we find evidence of previous exposure to ZIKV, we also found evidence of recent exposure (i.e. IgM seropositivity) to all three arboviruses. While we cannot rule out the potential for cross-reactivity influencing our results, they are suggestive of a large burden of *Ae. aegypti*-borne diseases in a setting where this important vector of disease is known to be present. Repeated infections can in some cases lead to more severe disease outcomes, for example through ADE. More research into arboviral diseases and their mosquito vectors in this setting is warranted both to confirm our results and to develop a better understanding of the burden of different arboviral diseases, any potential age and gender associated risk factors, and all so that public health interventions can be formulated to address these diseases. As the world has experienced pandemic levels of *Aedes*-borne disease several times since 2019, it is increasingly important to have increased surveillance for these diseases and for public health agencies to plan accordingly.

## Data Availability

All data produced in the present study are available upon reasonable request to the authors

## ACKNOWLEDGMENTS

We are thankful for the community members and community health workers in Dire Dawa and Addis Ababa who participated in this research.

