## Supplemental Materials for "High Seroprevalence to *Aedes*-borne arboviruses in Ethiopia: a Cross-sectional Survey in 2024"

**Supplemental Figure 1:** Age specific seroprevalence by arbovirus and for IgG (top panel) and IgM (bottom panel). Bar size is the proportion who tested positive ("reactive") by arbovirus. The

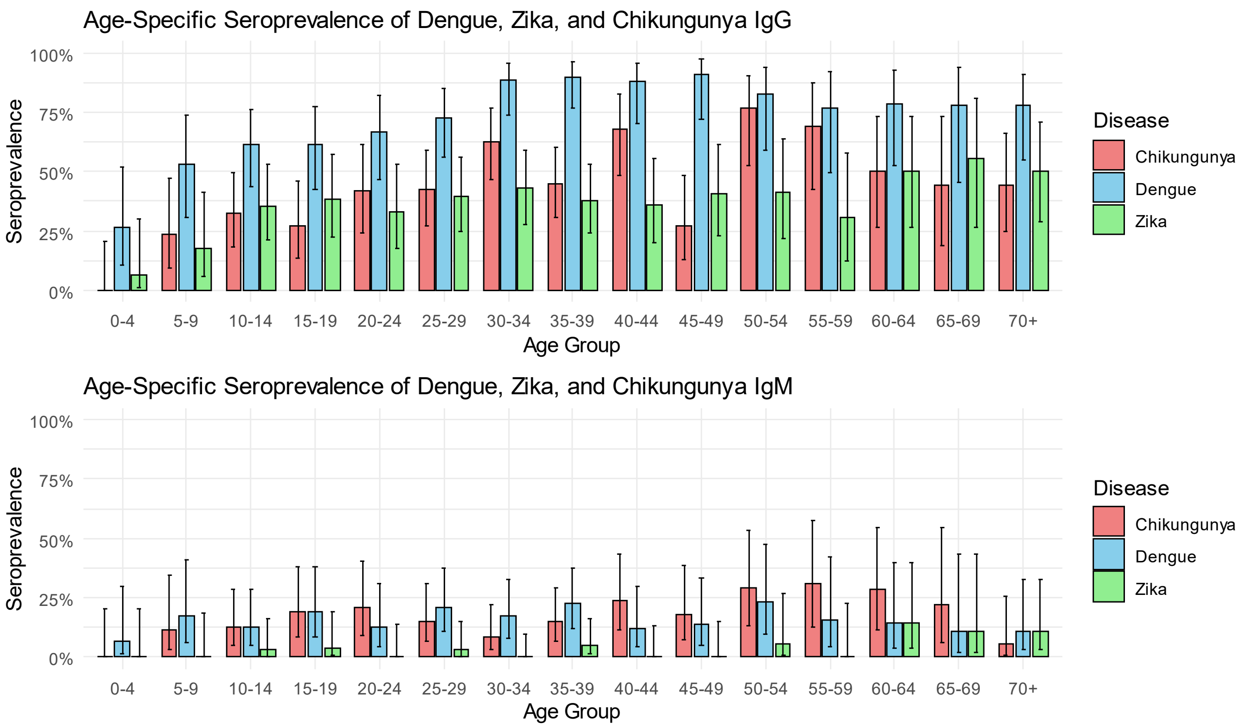


corresponding whiskers are Wilson score confidence intervals.

Age specific seropositivity for dengue IgG was high across most age groups, beginning at around 25% in the 0 to 4 age group and reaching a high of approximately 90% in the 30 – 34 age group. Chikungunya IgG seropositivity was next highest, reaching a high of 75% in the 50 – 54 age group. As with both dengue and chikungunya, Zika IgG seropositivity was lowest in the 0 – 4 age group, increasing with age but leveling off around age 15 – 25. All age groups exhibited IgG seropositivity to all three viruses.

IgM to dengue virus and chikungunya virus was higher than for Zika virus, though all three were present in almost all age groups (with an exception of Zika IgM being absent in the 0 – 4, 20 – 24,

30 – 34, and 40 – 49 age groups). Dengue IgM seropositivity was dominant in age groups up until the 40 – 45 age group, when chikungunya IgM seropositivity became highest.

**Supplemental Figure 2:**

IgG responses for ZIKV and DENV by test status. Since all ZIKV IgG positives were also DENV IgG positive, there is no way to look at the IgG among ZIKV IgG positives but DENV IgG negative (panel A). Panel B shows, for all individuals who were ZIKV IgG negative, the differences in quantitative ZIKV IgG response among those who were DENG IgG positive (blue/green) and those who were DENG IgG negative (orange). Panel C shows, for those who were DENG IgG positive, the difference in quantitative DENG IgG values for those who were ZIKV IgG positive or negative. DENV IgG was much higher among individuals who tested positive for ZIKV IgG (panel C). ZIKV IgG is also higher among those who tested positive for DENV IgG (panel B), but the difference is small in comparison to association between

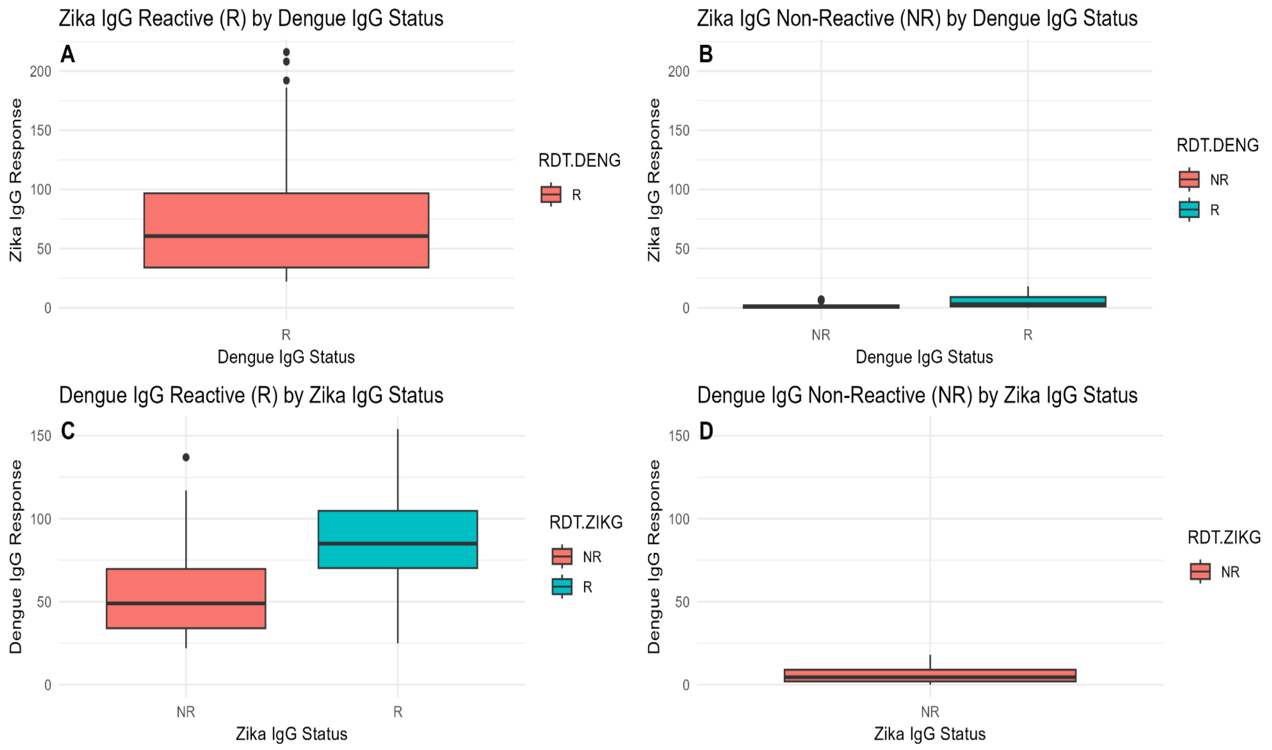

**Supplemental Table 1: Linear mixed models (random intercept for household) for IgG.** The "Model" column indicates which of the three viruses was the outcome while the "Predictor" column indicates the virus IgG measures that were used as predictors for in that model.

| Model | Predictor | Estimate | CI | Std.Error | p-value |
|---|---|---|---|---|---|
| Dengue IgG | (Intercept) | 32.10 | (27.47, 36.73) | 2.36 | <0.0001 |
| Dengue IgG | Zika IgG | 0.52 | (0.45, 0.59) | 0.04 | <0.0001 |
| Dengue IgG | Chikungunya IgG | 0.14 | (0.09, 0.18) | 0.02 | <0.0001 |
| Zika IgG | (Intercept) | -5.61 | (-12.11, 0.89) | 3.31 | 0.0915 |
| Zika IgG | Dengue IgG | 0.72 | (0.62, 0.82) | 0.05 | <0.0001 |
| Zika IgG | Chikungunya IgG | -0.07 | (-0.13, -0.02) | 0.03 | 0.0087 |
| Chikungunya IgG | (Intercept) | 23.85 | (11.20, 36.50) | 6.46 | 0.0003 |
| Chikungunya IgG | Zika IgG | 0.70 | (0.47, 0.93) | 0.12 | <0.0001 |
| Chikungunya IgG | Dengue IgG | -0.27 | (-0.48, -0.07) | 0.10 | 0.0084 |

**Supplemental Table 2: Linear mixed models (random intercept for household) for IgM.** The "Model" column indicates which of the three viruses was the outcome while the "Predictor" column indicates the virus IgG measures that were used as predictors for in that model.

| Model | Predictor | Estimate | CI | Std.Error | p-value |
|---|---|---|---|---|---|
| Dengue IgM | (Intercept) | 7.99 | (6.52, 9.47) | 0.75 | <0.0001 |
| Dengue IgM | Zika IgM | 0.33 | (0.19, 0.47) | 0.07 | <0.0001 |
| Dengue IgM | Chikungunya IgM | 0.39 | (0.29, 0.49) | 0.05 | <0.0001 |
| Dengue IgM | ZIKa IgG | 0.06 | (0.04, 0.09) | 0.01 | <0.0001 |
| Dengue IgM | Chikungunya IgG | -0.06 | (-0.08, -0.03) | 0.01 | <0.0001 |
| Zika IgM | (Intercept) | 0.09 | (-1.39, 1.57) | 0.76 | 0.9050 |
| Zika IgM | Dengue IgM | 0.24 | (0.16, 0.32) | 0.04 | <0.0001 |
| Zika IgM | Chikungunya IgM | 0.18 | (0.10, 0.26) | 0.04 | <0.0001 |
| Zika IgM | Dengue IgG | 0.01 | (-0.01, 0.04) | 0.01 | 0.2430 |
| Zika IgM | Chikungunya IgG | -0.03 | (-0.05, -0.01) | 0.01 | 0.0013 |
| Chikungunya IgM | (Intercept) | 3.43 | (0.39, 6.47) | 1.55 | 0.0281 |
| Chikungunya IgM | Dengue IgM | 0.42 | (0.27, 0.58) | 0.08 | <0.0001 |
| Chikungunya IgM | Zika IgM | 0.41 | (0.20, 0.62) | 0.11 | 0.0002 |
| Chikungunya IgM | Dengue IgG | 0.06 | (0.01, 0.12) | 0.03 | 0.0178 |
| Chikungunya IgM | Zika IgG | -0.08 | (-0.12, -0.03) | 0.02 | 0.0013 |
